# Multicenter Analysis on Morbidity, Mortality, and Medical Care in Neonates at the Hospital Gineco Obstétrico Isidro Ayora, Hospital General Docente de Calderón, and Hospital Gineco Obstétrico Pediátrico de Nueva Aurora Luz Elena Arismendi, January 2008 - June 2024, Quito-Ecuador (clinical research design protocol)

**DOI:** 10.1101/2024.06.09.24307243

**Authors:** Santiago Vasco-Morales, Mercedes Elina Yánez Valencia, Paola Toapanta-Pinta

**Affiliations:** Hospital Gineco Obstétrico Isidro Ayora; Hospital General Docente de Calderón; Hospital Gineco Obstétrico Pediátrico de Nueva Aurora Luz Elena Arismendi

## Abstract

**Introduction:** In 1983, the Latin American Center for Perinatology/Women’s Health and Reproductive Health published the Perinatal Information System, which records perinatal information of the mother and newborn. At the Isidro Ayora Gynecological Obstetric Hospital, there has been an electronic record of this database since 2008. In 2016, the Ministry of Public Health adapted and added registration variables to this medical history. Similarly, at the General Teaching Hospital of Calderón and the Pediatric Gynecological Obstetric Hospital of Nueva Aurora Luz Elena Arismendi, there are records of neonatal care such as neonatal anthropometry, perinatal risk factors, admission diagnoses, and length of hospital stay. These variables can be integrated with the database of the Isidro Ayora Gynecological Obstetric Hospital.

**Objectives:** Analyze the variables of the perinatal clinical history to identify factors associated with changes in fetal-neonatal morbidity and mortality.

**Methods:** Descriptive, analytical, observational study using secondary sources. Frequentist and Bayesian statistical analyses will be applied. To determine the association between qualitative variables, the Chi-square test and logistic regression models will be used. The t-test and linear regression will be used to analyze numerical variables. Statistical significance will be set at p<0.05, and Odds Ratios with a 95% confidence interval will be calculated. Neonatal growth curves stratified by sex and gestational age will be developed. The statistical program R will be used.

**Expected Results:** The characteristics of the population served in the hospitals in the north, central, and south of Quito, as well as the demographic and obstetric data of the mothers and their neonates, will be known. Perinatal characteristics associated with an increase or decrease in neonatal morbidity and mortality will be identified. Trends in maternal and child health will be detected and compared with national and international standards.

## Introduction

Care for mothers and their newborns is fundamental in public health and a research priority in Ecuador and worldwide. (Ministerio de Salud Pública del Ecuador, 2017; Yoneda et al., 2021)

According to the Ministry of Public Health, the main causes of neonatal mortality in Ecuador during 2020 were related to prematurity (26.13%), respiratory distress syndrome (18.14%), congenital malformations (17.28%), bacterial sepsis (11.66%), severe neonatal asphyxia (5.40%), pneumonia (2.86%), fetuses affected by maternal causes (1.73%), hemorrhages (1.51%), intrauterine hypoxia (0.65%), and sudden death (0.43%). During the same period, the Ministry of Public Health also reported that the Hospital Gineco Obstétrico Isidro Ayora ranked second among hospitals with the highest percentage of neonatal deaths, notified at 18.37%.^[3]^

The Centro Latinoamericano de Perinatología/Salud de la Mujer y Reproductiva (CLAP/SMR) is a technical unit of the Pan American Health Organization (PAHO) that provides technical assistance on sexual and reproductive health to countries in Latin America and the Caribbean. In 1983, the CLAP/SMR developed the Perinatal Information System (SIP), which comprises a set of tools designed for use in gynecology, obstetrics, and neonatology services. The perinatal clinical history is a detailed record initiated during pregnancy and extending to the postpartum period, collecting medical and obstetric data of the mother and newborn, along with procedures and treatments during pregnancy, childbirth, and postpartum. The SIP facilitates data collection into a database created through its software program, enabling the generation of local, national, or regional reports. These databases can be collectively analyzed to describe the evolution of various indicators over time, geographical areas, service networks, or population groups. All clinical information of the perinatal process is compiled on a single page, including additional forms for recording neonatal care if hospitalization is required. (Fescina et al., 2011)

The Ministerio de Salud Pública del Ecuador (MSP), as the governing body of the National Health System, adapted the Historia Clínica Perinatal from the Perinatal Information System (SIP) in 2016 to develop the Maternal Perinatal Clinical History (HCMP) “Form 051”. This update included variables related to maternal morbidity, severe maternal morbidity, interventions during childbirth, and the registration of the kangaroo method as treatment for premature or low-weight newborns. ^[5]^ Additionally, the option for Human Milk was added to the neonatal feeding record, corresponding to the administration of pasteurized maternal milk from a “Human Milk Bank”. The Hospital Gineco Obstétrico Isidro Ayora (HGOIA) has electronically registered the SIP Perinatal Clinical History database since 2008 and the HCMP since 2016. The Hospital General Docente de Calderón (HGDC) and the Hospital Gineco Obstétrico Pediátrico de Nueva Aurora Luz Elena Arismendi (HGONA) have records of neonatal care, including neonatal anthropometry, perinatal risk factors, diagnoses at hospital admission, and hospital stay time. These databases enable the creation of a database that will allow us to know the characteristics of the served population, evaluate the outcomes of the care provided, and identify the risk factors associated with neonatal morbidity and mortality in Quito.

According to the Ministerio de Salud Publica (MSP), the main causes of neonatal mortality in Ecuador during 2020 were related to prematurity (26.13%), respiratory distress syndrome (18.14%), congenital malformations (17.28%), bacterial sepsis (11.66%), severe neonatal asphyxia (5.40%), pneumonia (2.86%), fetuses affected by maternal causes (1.73%), hemorrhages (1.51%), intrauterine hypoxia (0.65%), and sudden death (0.43%). During the same period, the MSP also reported that the HGOIA ranked second among hospitals with the highest percentage of neonatal deaths, notified at 18.37%. ^[3]^ It should be considered that the HGOIA is a tertiary care hospital and a national referral center, so patients with high-risk pregnancies and fetuses with complex pathologies are referred to this institution. However, data from the Hospital General Docente de Calderón (HGDC) and the Hospital Gíneco Obstétrico Pediátrico de Nueva Aurora Luz Elena Arismendi (HGONA) can complement the study of perinatal characteristics in the city of Quito.

Below are the medical conditions that will be analyzed in this project.

Perinatal hypoxic-ischemic disorders are a serious health issue that can lead to long-term brain damage and disability. Among these disorders, perinatal asphyxia stands out, causing 2.5 million neonatal deaths globally and contributing to 47% of deaths in children under 5 years old. Maternal and neonatal risk factors have been identified to date; however, effectively preventing these disorders remains a challenge. Early diagnosis, proper identification of specific risk factors for each population, and timely treatment are crucial in reducing neonatal morbidity and mortality associated with hypoxic-ischemic perinatal pathology. (Moshiro et al., 2019)

Congenital malformations are significant causes of infant mortality, chronic disease, and disability, according to the World Health Organization (WHO), approximately three hundred thousand children die worldwide due to congenital anomalies.^[7]^ These congenital malformations are often associated with chromosomal defects, which can be caused by a combination of genetic and environmental factors; a variety of risk factors have been identified, including advanced maternal age, exposure to teratogens during pregnancy, chronic maternal diseases and those specific to pregnancy, consanguinity, and a family history of congenital defects. Additionally, advances have been made in identifying and characterizing specific genes associated with certain congenital disorders.^[7–9]^ However, there are pathologies that are more common in certain populations, therefore, a detailed characterization of congenital malformations presented in neonates born at the Hospital Gineco Obstétrico Isidro Ayora (HGOIA), Hospital General Docente de Calderón (HGDC), and Hospital Gineco Obstétrico Pediátrico de Nueva Aurora Luz Elena Arismendi (HGONA), along with their risk factors and demographic characteristics, is necessary.

Neonatal infections and their most severe complication, such as neonatal sepsis, are significant and preventable causes of neonatal morbidity and mortality. A study conducted in 2017 identified 5.1 million cases of neonatal sepsis worldwide. (Rudd et al., 2020) These infections are caused by bacteria, viruses, and parasites that can be transmitted from the mother to the fetus during pregnancy, childbirth, and the postpartum period. Premature neonates and those with low birth weight are at higher risk due to their immature immune systems, making them more susceptible to infections. Other risk factors include prolonged rupture of membranes (>18 hours), maternal fever during childbirth, maternal colonization with pathogenic microorganisms, and maternal infections such as HIV, syphilis, herpesvirus, or other microorganisms.^[11]^ To prevent neonatal infections, timely treatment of maternal infections is recommended, along with applying prophylactic measures against infections, such as administering antibiotics to prevent Group B Streptococcus and immunizations. Additionally, it is important to emphasize the rational use of antibiotics during childbirth and neonatal hospitalization. However, prevention is one of the best strategies, and within this process, the identification of associated risk factors is essential. (Good & Hooven, 2019)

The causes of fetal and neonatal death are closely related to preterm birth, low birth weight, preeclampsia, gestational diabetes, maternal infection, smoking, alcohol and drug consumption, and advanced maternal age as the main factors. In Ecuador, there are still many uncertainties surrounding the prevention of fetal-neonatal death and the level of influence of associated perinatal risk factors, which are not precisely known. (Richtmann et al., 2020; Zamaniyan et al., 2020)

According to the World Health Organization, 15 million premature babies are born every year, and the chronic complications arising from prematurity represent a global public health problem. Among the identified risk factors, birth weight, gestational age, exposure to infections during pregnancy, maternal nutrition, and neonatal medical interventions such as mechanical ventilation or prolonged hospitalization stand out. In this regard, premature newborns are at a higher risk of developing chronic diseases in the medium term, such as bronchopulmonary dysplasia, neurological and neurodevelopmental disorders, and retinopathy of prematurity. Prematurity can also have long-term consequences, including cardiovascular, metabolic, psychological, and socioeconomic diseases, affecting both children and their families. (OMS, 2020; Ortega Barrionuevo & Vasco, 2022; World Health Organization, 2023)

Intrauterine growth disorders such as low birth weight (less than 2500 grams) and macrosomia (weight over 4000 grams or above the 90th percentile, according to growth curves for gestational age) can have consequences for the health of the newborn, including a higher risk of neonatal mortality, disability, and long-term chronic diseases. Globally, it is estimated that between 15 and 20% of newborns are born with low birth weight. (Amigo et al., 2015; Bashinsky, 2017; M et al., 2013) According to the Ecuadorian Institute of Statistics and Census (INEC), in 2021, 8.6% of children were born with low birth weight. ^[21]^ Regarding fetal macrosomia, in the United States of America, 7.8% of neonates are reported to be macrosomic. In Ecuador, proportions range from 3.8% to 11.3%, with the latter value corresponding to a sample collected at the Isidro Ayora Gynecological-Obstetric Hospital (HGOIA) and hospitalized patients only.^[22–24]^ Both issues are associated with maternal risk factors such as age, nutrition, tobacco and alcohol consumption, and the presence of chronic diseases. It is not clear which risk factors are most associated with these disorders in our population. Therefore, further research is needed to better understand the risk factors associated with intrauterine growth disturbances and to develop more effective strategies to prevent their occurrence.

The health programs integrated into the Maternal-Perinatal Clinical History (HCMP), implemented to enhance neonatal health at HGOIA, HGDC, and HGONA, include the Kangaroo Mother Care program ESAMyN and the distribution of breast milk through a human milk bank, aimed at feeding sick neonates.^[25,26]^ Currently, human milk banks are widespread in hospitals and neonatal care units, where donated breast milk is collected, processed, stored, and dispensed for use in feeding premature neonates or those with other special health conditions. In 2007, HGOIA inaugurated Ecuador’s first human milk bank. Presently, some studies have investigated maternal and neonatal conditions associated with attendance at the human milk bank, such as gestational age at birth, neonatal birth weight, maternal age along with educational level, type of delivery, received social support, breastfeeding counseling, variables included in the HCMP. It has been demonstrated that feeding with either own or donated breast milk provides a range of benefits for neonates, including reducing infection rates, decreasing hospitalization days, and overall neonatal morbidity and mortality. (MSP, 2016)

At HGOIA, the Kangaroo Mother Care method was implemented in 1991 as part of a pioneering study that showcased the benefits of this practice to the world.^[27]^ This method is an effective strategy for improving the morbidity and mortality rates of premature and low birth weight newborns, and it is currently also being applied at HGDC and HGONA. Maternal conditions such as age, educational level, access to healthcare services, mental health, and partner involvement have been found to influence a mother’s willingness to adopt the Kangaroo Mother Care method. On the other hand, neonatal conditions like gestational age, birth weight, need for mechanical ventilation, and the presence of respiratory and cardiac diseases can impact a newborn’s ability to adapt to this method. The successful implementation of Kangaroo Mother Care also relies on the availability of resources and support from healthcare personnel, the quality of care, and hospital infrastructure. (Kostandy & Ludington-Hoe, 2019) Ultimately, monitoring, and early identification of perinatal conditions that may pose as risk factors or protective factors are variables that can be studied in both the SIP and the HCMP of hospitals participating in this multicenter study. The results of this study will enable the implementation of preventive and therapeutic interventions to reduce neonatal and fetal morbidity and mortality in Quito.

### General Objective

To analyze the variables of the perinatal clinical history to identify factors associated with changes in fetal and neonatal morbidity and mortality in patients treated at HGOIA, HGDC, and HGONA.

## Specific Objectives

1. To identify risk factors associated with the main causes of neonatal and fetal morbidity and mortality: hypoxic-ischemic disorders, congenital malformations, neonatal infections, and prematurity.
2. To identify risk factors associated with intrauterine growth disorders: low birth weight and macrosomia.
3. To identify risk factors associated with complications of prematurity, such as bronchopulmonary dysplasia, retinopathy of prematurity, and neurodevelopmental disorders.
4. To identify maternal and neonatal conditions related to adherence to the Kangaroo Mother Care plan and attendance of mothers at the Human Milk Bank.

### Study Design

Cross-sectional study with retrospective data collection.

### Population Definition

From January 2008 to December 2023, 116,603 births have been recorded at HGOIA. For this study, the plan is to include the entire population in HGOIA, while in HGDC and HGONA, the plan is to include all neonatal population born since their inauguration in 2015 and 2016 respectively.

### Data Collection

Data will be obtained from neonates in the three hospitals during the specified period, considering the inauguration dates of HGDC and HGONA.

### Data Cleaning

Review the data to identify any errors, inconsistencies, or missing information. Clean the data by removing duplicate entries, detecting errors, and addressing missing information where possible.

Identifying the risk factors associated with the study variables.

Bivariate and multivariate statistical analyses will be conducted to determine the strength of association between maternal and neonatal variables related to each identified risk factor according to the pathology or group of pathologies studied, by calculating OR and their 95% CI. Regression models will be adjusted based on birth weight or gestational age as appropriate. Indicators will be compared among the three hospitals to identify significant differences in neonatal morbidity and medical care. This comparison will consider the characteristics of the patient population, level of complexity, and other factors identified as influential in the outcomes and observed differences between hospitals.

Drafting at least three scientific articles will result from the compilation, exhaustive data analysis, and comparison of the main results with relevant publications on each topic. This will help identify the risk factors associated with causes of neonatal morbidity, characterize detected congenital malformations and their associated risk factors, and finally, explore the causes and risk factors associated with fetal and neonatal mortality.

### Statistical Analysis Software

To analyze the extracted data and generate graphs, models, etc., the program R will be used in this case.

Maternal characteristics of interest were age, educational level, and ethnic group. According to the national census, there are five major ethnic groups in Ecuador: Mestizo, Afro-Ecuadorian, Indigenous, Blanco (White). Mestizos are historically defined as the descendants of Europeans and Indigenous Americans, and constitute the largest part of the Ecuadorian population. Afro-Ecuadorians have high fractions of African and Native American ancestry. Information about maternal obesity (WHO International Classification of Diseases 10th revision, ICD-10, code E66), gestational hypertension (ICD-10 code O14), gestational diabetes (ICD-10 codeO24), premature rupture of membranes (PRM, ICD-10 code O42), and anemia complicating pregnancy (ICD-10 code O990) will be obtained from the mothers’ medical records. Delivery characteristics of interest were the newborn’s sex, position during labor, birth weight, and gestational age.

Prepartum risk was defined as imminent, high, or low risk, according to the classification determined by the IMCI (Integrated Management of Childhood Illnesses) strategy. A gestation is classified as at imminent risk if at least one of the following signs is present: pregnancy >41 weeks, decreased or absent fetal movements, severe systemic disease, urinary tract infection with fever, uncontrolled diabetes, uncontrolled hypertension and/or presence of seizures, blurred vision, loss of consciousness or severe headache, severe paleness and/or hemoglobin (Hb) <7 g/dl, edema of the face, hands and legs, or PRM before 37 weeks. Gestation is classified as high risk if it presents at least one of the following signs: maternal age <20 years or >35 years, primiparous or large multiparous, period between pregnancies less than two years, no prenatal control, uterine height does not correlate with gestational age, history of preterm, low birth weight or malformed children, history of habitual abortion, early fetal or neonatal death, controlled systemic disease, urinary tract infection without fever, controlled diabetes, controlled hypertension, palmar pallor and/or Hb ≥7 and <12 g/dl, refractory vaginal discharge, alcohol, tobacco or drug abuse, history of violence or abuse, teratogenic drug intake, inadequate weight gain, body mass index <20 or >30 kg/m2, abnormal presentation, multiple gestation, Rh-negative mother, VDRL or HIV or hepatitis B positive, oral or periodontal problems, or no tetanus toxoid immunization. A low-risk gestation is one that is classified neither as high risk nor as imminent risk.

Low birth weight (LBW) is defined by the WHO as a birthweight of less than 2500 g (5.5 lb), regardless of gestational age. Newborns were classified according to their birth weight as small (SGA), appropriate (AGA), and large-for-gestational-age (LGA). SGA was defined as a birthweight below the 10th centile birth weight for gestational age and sex with the INTERGROWTH-21st birth weight standard, and LGA was defined as a birthweight >90th centile

Prematurity was defined as a gestational age of less than 37 weeks, and it is a leading cause of infant mortality, morbidity, and long-term disability. The WHO subdivides the preterm births into three groups based on the gestational age: extremely preterm (<28 weeks), very preterm (28 to <32 weeks), and moderate or late preterm (32 to <37 completed weeks of gestation) (World Health Organization, 2012). Infants born below 34 weeks experienced substantial morbidity and often required admission to neonatal intensive care units.

### Data Storage

To store the extracted data and the intermediate or final analysis results. The anonymized database will be stored on the principal investigator’s personal computer hard drive, electronically in Mendeley Data, with restrictions on public download.

## Ethical and Gender Considerations

The management of neonatal information pertains to a vulnerable population; therefore, in this research, like others involving human subjects, it will be guided by ethical principles ensuring respect for individuals and the community. These principles include respect for autonomy and voluntary participation, with a particular focus on ensuring the protection of data privacy and confidentiality in this study. Furthermore, being a secondary source study, it is considered a minimal risk study where the benefits outweigh the risks, as this research could have a significant impact on patient care at HGOIA, HGONA, and HGDC. The results of this research will be published in indexed journals.

## Expected Results

### OE1

Prevalence, description, and risk factors associated with the main causes of neonatal and fetal morbidity and mortality.

### OE2

Prevalence, description, and risk factors associated with intrauterine growth disorders: low birth weight and macrosomia.

### OE3

Prevalence, description, and risk factors associated with complications of prematurity, such as bronchopulmonary dysplasia, retinopathy of prematurity, and neurodevelopmental disorders.

### OE4

Prevalence, description, and identification of maternal and neonatal conditions that favor adherence to the Kangaroo Mother Care plan and attendance of mothers at the Human Milk Bank.

The obtained information will be relevant for identifying risk factors, evaluating medical interventions, and making clinical decisions for the treatment of hospitalized neonates in the neonatology services of HGOIA, HGONA, and HGDC. Additionally, the results may be applicable in improving prenatal care, reducing fetal and neonatal morbidity and mortality, and drafting treatment protocols and guidelines to be implemented in the mentioned hospitals.

There are several limitations and biases that may prevent obtaining precise and reliable results when analyzing the perinatal clinical history from the SIP and HCMP. Some of these limitations and biases include:

Errors in clinical history documentation: There may be errors, incomplete data, or inconsistencies recorded in the perinatal clinical history, making it difficult to obtain accurate information.

Information biases: Patients may have provided incomplete or incorrect information due to memory failure or the influence of external factors, such as situations where patients arrived at the emergency department in critical condition.

To reduce these biases, a thorough evaluation of the data will be conducted to only include the population subset with complete and accurate data.

## Data Availability

No data

